# Evaluating the impact of age on prostate cancer overdiagnosis using long-term follow-up from a randomised trial

**DOI:** 10.64898/2026.01.26.26344830

**Authors:** Adam R Brentnall, Matejka Rebolj, Peter Sasieni, Garth Funston, Rhian Gabe, Andrew Vickers

## Abstract

Prostate cancer overdiagnosis is detection of prostate cancer through PSA testing that otherwise would not have been diagnosed within the patient’s lifetime. It is a major concern to policymakers due to its impact on quality of life. We used long-term followup data from the CAP randomised trial of a one-off screen, and English male competing mortality rates (2021-23), to estimate the impact of age on excess prostate cancer incidence within 15 years (‘overdiagnosis’) using competing risks methods. In total, 2249 (1.19%) of 189,386 men invited for a PSA test in CAP had cancer detected at the one-off screen. Prostate cancer cumulative incidence at 15 years was 7.08% (95%CI 6.95 to 7.21%) in those invited to screening, compared with 6.94% (95%CI 6.82 to 7.06%) in the control arm; an absolute excess incidence difference of 0.14% (95%CI -0.04% to 0.37%). Excess net incidence to 15 years was 0.14/1.19 = 11.7% (95%CI 0.0% to 26.7%) of cases detected at a single prevalent screen. Accounting for competing mortality, English men diagnosed aged 50 years were projected to have a 16% chance the cancer would not have been detected within 15 years, rising to 32% aged 70 years and 58% aged 80 years. Thus, prostate cancer overdiagnosis rises substantially with age due to competing mortality, and is relatively low for younger men. Accordingly, opportunistic testing policies should be re-examined in settings where they have led to high rates of screening in older men.

## 1. Introduction

The evidence that prostate-specific antigen (PSA) screening reduces prostate cancer mortality has become clearer with extended follow-up from two large randomised trials.[1, 2] However, there remains considerable uncertainty whether the benefits of PSA screening, in terms of reduced mortality, outweigh the harms from PSA testing, in particular overdiagnosis. While some guideline developers recommend risk-based PSA screening,[3] many high-income countries, including the UK, have not recommended population-based PSA screening, largely on the grounds of the harms estimated from overdiagnosis.[4] Prostate cancer overdiagnosis is detection of prostate cancer through PSA testing that otherwise would not have been diagnosed within the man’s lifetime. It occurs when a man, or other person with a prostate, has prostate cancer detected from an opportunistic or organised programme screening, but dies from other causes before the cancer would have presented clinically following symptoms without screening. The main harms due to overdiagnosis are from subsequent disease management that negatively impacts quality of life. Men with overdiagnosed prostate cancer have an increased risk of, among others, incontinence, erectile dysfunction and bowel irritation, and anxiety if the prostate cancer is monitored for signs of progression. The risk of harm in men screened has been reduced but not eliminated through the use of magnetic resonance imaging (MRI) in the diagnostic pathway[5]; approximately 8% of men diagnosed with low-risk cancer in England and Wales nonetheless received radical treatment in 2021.[6]

Accurate evaluation of overdiagnosis is difficult. In theory overdiagnosis could be observed directly in a study where men had PSA tests and diagnostic workup but were blinded to whether they had prostate cancer detected. Men who subsequently died without a prostate cancer diagnosis over the rest of their life, but who had it detected at screening, would have been overdiagnosed by screening. Of course, such a study design would be unethical and impractical and so evaluation of overdiagnosis requires indirect methods. One method used is modeling, including microsimulation approaches where the natural history of prostate cancer is simulated in a cohort of digital humans. This approach was done to inform prostate cancer screening recommendations by the UK National Screening Committee.[7] A difficulty of such models is that they require many assumptions. Any approach that depends on a large number of assumptions, many of which cannot be tested, needs to be shown to be well calibrated to empirical data before projections are accepted. For instance, when the model is used to compare a new policy with current standard of care, the model should match epidemiological parameters such as incidence and mortality and other key aspects of the harm/benefit equation such as overdiagnosis. To help evaluate the plausibility of such complex models, we aimed to estimate overdiagnosis by age using data on the excess of cancers that would be diagnosed over a person’s lifetime as a result of screening, vs no screening. For this we used data from a mature randomized trial with a stop-screen design and long follow-up after screening.[1] This has previously been considered the gold standard approach to estimate overdiagnosis.[8] While the screening pathway for prostate cancer has changed since the trial was run, it is unlikely that we will have better data to estimate overdiagnosis with modern pathways using the excess incidence approach from other studies anytime soon. Therefore, we need to make the best use of such data to inform policy now, and we end by considering potential implications for policymakers from our analysis.

## 2. Materials and methods

The UK Cluster Randomized Trial of PSA Testing for Prostate Cancer (CAP) trial evaluated a one-off prostate cancer PSA test.[1] Between 2001-2007, n=195,912 men were cluster randomised to a one-off invitation for a PSA screening test, and n=291,445 men to control. A screen-positive test result was indicated if PSA was above 3.0 ng/mL. Ten-core transrectal ultrasonography-guided biopsies were used for further investigations. Follow-up data on vital status and cancer registration through December 21, 2020 were from National Health Service databases. Follow-up data were complete for 97% randomized to the intervention and 99% to control, up to December 31, 2020, with a median of 15 years (IQR: 14.2-16.4; range, 12.2-19.2) after randomisation. The trial took place in a setting and era when there was relatively little opportunistic PSA testing in the target population. n=64,425 (34%) men randomised to screening had a valid test result, and cumulative PSA testing in the control arm was estimated by the trialists to be 10-15% over the first 10 years.[1]

We evaluated one of the epidemiological definitions of overdiagnosis using excess cumulative incidence *O*(*t*) to *t* years after a one-off screen, considering the absolute difference in risk *P*_*A*_(*t*) − *P*_*B*_(*t*) between those randomised to screening arm *A* vs control arm *B*, relative to the absolute risk of screen-detected cancers at the one-off screen *R*_*A*_:

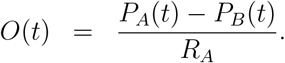

A long follow-up time *t* = *T* is needed for reliable estimation of overdiagnosis.[9] The median 15-year follow-up in CAP is substantially longer than other prostate cancer screening trials with (repeat) PSA testing.[1] For shorter follow-up, the formula for *O*(*t*) does not measure overdiagnosis but observed cumulative excess to a given time *t*. This need not correspond to overdiagnosis if the expected ‘compensatory drop’ in incidence in the screening arm has not ended, so that the excess could decrease further beyond the latest follow-up. The compensatory drop in incidence is expected once screening stops, because some cancers that would have been diagnosed then were diagnosed early from screening. With *n*_*A*_ : *n*_*B*_ randomisation and complete follow-up to *T* years after screening (no censoring other than death), overdiagnosis may be estimated using the observed cumulative number of cancers *Y*_*j*_(*T*) through *P*_*j*_(*T*) = *Y*_*j*_(*T*)*/n*_*j*_ for *j* = *A, B*, with *R*_*A*_ as the percentage of those randomised with screen-detected cancer at the one-off screen. An issue to contend with is that subjects are sometimes are censored before time *T* in a trial. This might occur for several reasons, including randomisation taking place over an extended period of calendar time, loss to follow-up, or death. If the censoring process is independent of prostate cancer incidence, then one may estimate net risk where the impact of the censoring process is removed in the standard way, by using the complement *F* = 1 − *S* of Kaplan-Meier estimates. This estimates the incidence of prostate cancer irrespective of censoring, and so in the CAP trial data, irrespective of death from other causes. One may then define ‘net’ excess *E*(*t*) at *t* years after screening using:

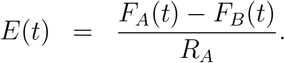

To estimate net excess from CAP we extracted Kaplan-Meier estimates of prostate cancer incidence by arm and follow-up time 0 < *t* ≤ 15 (Figure 2c of [1] using https://plotdigitizer.com/; our Figure 1a). Then to ensure overdiagnosis was not greater than 100%, we estimated the net excess at *t* as *E*(*t*) = min[1, {*F*_*A*_(*t*) − *F*_*B*_(*t*)}*/R*_*A*_]. We set *R*_*A*_ = 1.19% because it has been reported that 2249 (1.19%) of 189,386 men invited for a PSA test in CAP had cancer detected at the baseline screen.[10] To estimate overdiagnosis it is necessary to consider two competing risks: (1) net excess, and (2) competing mortality. For example, if everyone died within 1 year (eg. due to a natural disaster) then overdiagnosis would be 100%. If nobody died within 15 years (∼impossible) then net excess would equal crude excess. The truth will lie in between these extremes depending on rates of competing mortality, with overdiagnosis being higher for those with higher competing mortality rates such as older men. To proceed we assumed that net excess is independent of competing mortality. This is reasonable for prostate cancer, as it is not strongly linked with other causes of death. We further assumed net excess is independent from age; a weaker assumption than assuming that incidence is independent from age (see discussion for evaluation of risk of bias from this assumption). Then we applied standard competing-risks methodology, treating *E*(*t*) = *S*_1_(*t*) as a survival function (for the first competing risk).[11, 12] For the second competing risk, competing-mortality rates *h*_2_(*a*) by year of age *a* were taken from all-cause English male mortality rates 2021-23,[13] subtracting age-specific prostate-cancer mortality rates.[14] Annual excess rates were calculated as *h*_1_(*k*) = log{*S*_1_(*k* − 1)} − log{*S*_1_(*k*)} by year since randomisation *k* = 1, 2, …, 15. Then crude excess for a man aged *a* years, *t* years after screening, was from:

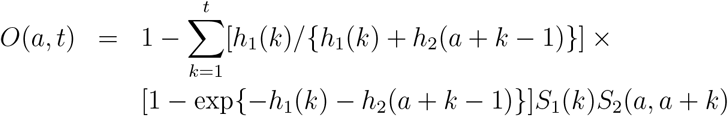

where 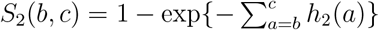. Overdiagnosis by age *a* was projected excess at 15 years {*O*(*a*, 15)}. Further analysis on the closing of the compensatory drop evaluated the hazard ratio for incidence from the trial as a function of follow-up time using the gradient of the derived cumulative hazard function, and a smoother. Confidence intervals from reported summary statistics were estimated using a z-statistic with standard errors (SE) from reported confidence intervals (95% confidence interval width divided by 2*1.96). All analysis was done in the statistical software R, and source code and data used are fully available.

**Figure 1:**
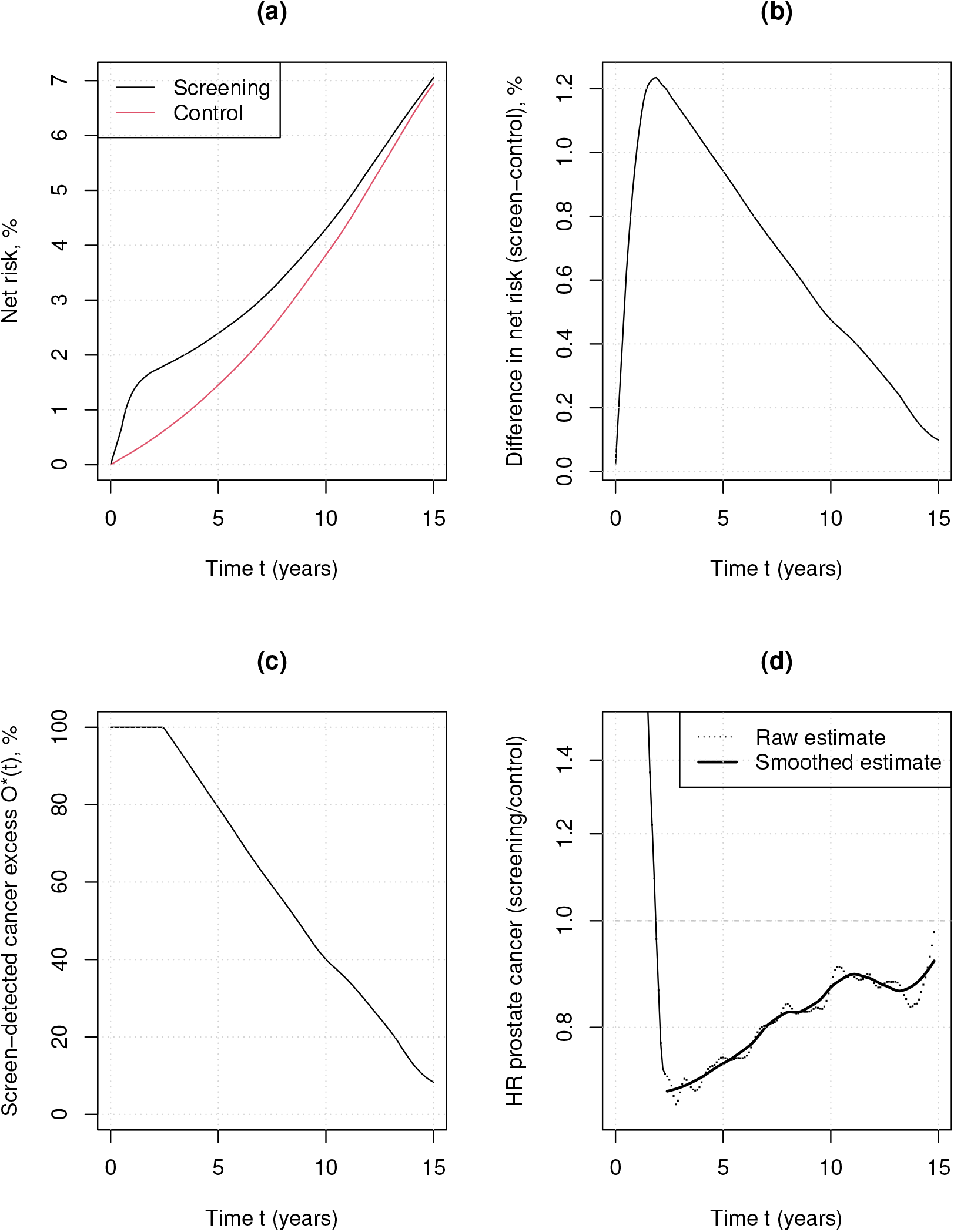
Estimates of net overdiagnosis and excess cancer incidence using data reported at 15 years follow-up from the CAP trial. Panel (a) shows the net risk estimates (Kaplan-Meier),[1] (b) is the absolute difference in net risk between the screening and control arm from panel (a); (c) is the observed excess *O*^∗^(*t*) as a function of follow-up time *t*, being the y-value from plot (b) divided by a baseline screen detection rate of (2,249/189,384),[10] capped at 100% during early follow-up, leading to an estimate of overdiagnosis at *t* = 15 years; (d) shows the ‘compensatory drop’ of cancer incidence following screening using an estimate of the hazard ratio (HR) for prostate cancer incidence as a function of follow-up time, based on data from (a).

## 3. Results

### 3.1. The impact of follow-up time on excess incidence

Prostate cancer net cumulative incidence was 7.08% (95%CI 6.95 to 7.21%) at a median 15 years in CAP in those invited to a one-off screen, compared with 6.94% (95%CI 6.82 to 7.06%) in control; a net excess incidence difference of 0.14% (95%CI -0.04% to 0.37%; Figure 1b).[1] Equivalently, relative net excess incidence of prostate cancer at 15 years was approximately (a statistically non-significant) 2% (7.06/6.94) higher. Since 1.19% of the screening arm were reported to have prostate cancer detected at the baseline screen,[10] net excess to 15 years was 0.14*/*1.19 = 11.7% (95%CI 0.0% to 26.7%) of cases detected at the single trial screen. The confidence interval is relatively narrow, being based on *>*24 thousand men with prostate cancer, and it includes 0%. The role of follow-up time on net excess incidence is illustrated in Figure 1c. Here the net excess with five years of follow-up was approximately 80% of screen-detected cases, so that at most approximately 20% of the cancers diagnosed at screening would have presented clinically in the first five years. This drops to 40% with ten years follow-up. Even with a median 15 years follow-up after screening data from CAP do not appear fully mature, and it remains to be seen if the compensatory drop in cancer incidence has ended (Figure 1d), ie. whether PSA testing can identify cancers with a lead time of more than 15 years.

### 3.2. The impact of age on overdiagnosis

The average English man aged 50 years has a 10% chance of death in the next 15 years, rising to 20%, 30%, 49% and 89% if they are respectively 59, 64, 70 or 80 years of age.[13] The ability of PSA testing to identify prostate cancer a decade or more early means that many more will be overdiagnosed due to death from other causes than prostate cancer, following PSA tests done at an older age. This can be quantified using Figure 1c, which suggests that the excess decreases from year 3 after screening by 7.36% per year {(100-11.7)/12} through to year 15. We used this assumption together with the competing mortality assumptions to estimate the proportion of screen-detected cancers that would have been diagnosed within 15 years by age, allowing for different rates of death from other causes. These calculations projected that the average man alive in England aged 50 years and diagnosed with prostate cancer at screening has a 16% chance to not have been diagnosed with prostate cancer within 15 years if he had not attended screening. If the maximum sojourn time is 15 years (which would conincide with closing of the compensatory drop), then this is an estimate of overdiagnosis (but if the maximum sojourn time is more than 15 years then this estimate of overdiagnosis will be too high). To illustrate the impact of age further, for men aged respectively 59, 64, 70 and 80 years of age at diagnosis following screening, overdiagnosis defined in this manner is estimated to be respectively 20%, 24%, 32%, and 58%. Note that competing mortality and hence overdiagnosis risk will be slightly lower for the average man currently receiving PSA testing in the UK due to other demographic factors than age, including comorbidities.[15]

## 4. Discussion

### 4.1. The main driver for overdiagnosis from PSA testing is age

We evaluated overdiagnosis from a one-off screen by using a competing risks analysis. We first estimated the net excess incidence from a large unique randomised trial with a one-off (stop) screen design and long, complete follow-up. To estimate overdiagnosis we combined this estimate with national data on competing mortality rates in English men. The data from CAP showed that net excess incidence from screening continues to decrease through to 15 years after screen detection and it is not clear if the trend has ended at 15 years. This long lead time means that overdiagnosis increases substantially with age of screen detection due to competing mortality. While the importance of age is consistent with the broader literature,[16] our overdiagnosis estimates are lower for younger men than some other studies. The main reason is that analysis considered a long 15-year follow-up after screening. ERSPC is the only other large randomised trial with complete recruitment that may generate suitable data to estimate overdiagnosis via the excess-incidence method. Excess incidence in the most recently published analysis from ERSPC remains higher than the CAP trial (crude cumulative incidence approximately 14% in the intervention group and 12% in the control group), but this is at least partly because the median follow-up time after last screen in the analysis was only 8 (inter-quartile range 5-12) years. For example, eight-year follow-up would only allow for up to half of screen-detected cancers at a prevalence screen to have presented symptomatically, even if the man did not die over that period (Figure 1c). Therefore, the expected compensatory drop of cancer incidence in the screening arm of the ERSPC trial has not yet fully materialised. The importance of extended follow-up has been seen in studies of overdiagnosis of lung and breast cancer.[9, 17] These analyses have shown that overdiagnosis estimates decrease with follow-up time, and that variation in estimates of overdiagnosis between studies is largely explained by follow-up time.

### 4.2. Limitations and generalisability

There are some reasons our estimates may be too low. The most important of these is there was contamination in CAP from opportunistic PSA testing (as in the other trials).[1, 18, 19] However, the trial was conducted and follow-up data were collected in a period when opportunistic testing (which likely took place in both arms) was relatively low. The control group were also not directly informed about the trial, whereas the intervention group all received some information about the trial and PSA testing. Therefore, we believe it is probable that the *differential* rate of asymptomatic testing subsequent to the screening is unlikely to change our main conclusions. Even if asymptomatic PSA testing only occured in the control group at the maximum level estimated by the CAP triallists (15%), and no further PSA testing occured at all to the initial 36% in the screening arm subsequently, overdiagnosis would not increase substantially if the estimate was adjusted upwards to reflect this. In a extreme situation where the net excess at 15 years is too low by a factor of (36-15)/36, then it would increase from 11.7% to 20.1%. Our estimates are also at risk of bias for repeat screening policies since they use data from a one-off screening intervention. Cancers detected at subsequent screening rounds are expected to be, on average, identified earlier in the natural history of prostate cancer than those at a first round. In other words, the estimate is at risk of spectrum effects, such that one would expect the average lead time to be longer for cancer detected at subsequent screens. This would serve to slightly increase excess incidence relative to our analysis, and longer follow-up would be needed than 15 years after last screen to reliably estimate overdiagnosis. Another potential issue is whether the results are generalisable to opportunistic testing in current practice. Opportunistic testing is screening but without organisation (including, eg., invitations to the entire eligible population at regular intervals). While there may be some (self) selection effects from opportunistic populations that are hard to anticipate, we believe the findings are likely directly relevant.

There are some reasons our estimates may be too high. The first is that excess incidence might further decrease beyond 15 years (Figure 1d), and we have only considered follow-up to 15 years after screening. Another is that the estimates effectively assume that no screen-detected cancers would present with symptoms within 3 years of screening, since the net excess is 100% for the first three years. Therefore, there is a risk of bias in the denominator taken for proportion of screen detected cancers (it might be too small). The analysis also treats the lead-time distribution to be the same by age, but other analyses have suggested that lead time may be less for younger men.[1] If true, overdiagnosis would be lower for younger men and even higher for older men. Lead time might also be shorter for men today because it is affected not only by age but also by biopsy protocols. Recent years have seen the development of further imaging and biomarker approaches that are used as secondary tests after a raised PSA to determine eligibility for biopsy. Randomized trials have shown that such approaches reduce detection of low-grade cancers most likely to result in overdiagnosis.[20, 5] Another potential bias is from the relatively low uptake of screening in the intervention arm. This might make the estimate of overdiagnosis too high because attenders in CAP were more likely to live in areas of lower deprivation,[21] and therefore to have previously had a PSA test and a higher risk of indolent disease than non-attenders.

In summary, our analysis is at risk of bias in different directions, but taken together we suggest that the analysis and interpretation is likely to be reasonable. Indeed, the time of ∼8 years to 50% excess, is consistent and no lower than other estimates of the average lead time from PSA testing in the pre-MRI era.[16]

### 4.3. Policy implications

There will be substantial overdiagnosis in men older than 70 years at screening, but much less overdiagnosis in younger men, eg. those aged 50-64 years. Screening older men is also unlikely to confer benefit in terms of mortality reduction. This was observed directly in the ERSPC trial where data suggested little benefit from screening men aged 70-74 years (95%CI 0.80 to 1.41).[22] This is because prostate cancer screening has a long lead time and there is relatively good survival from prostate cancer. Therefore, by the time a man could benefit from early screen detection older than 70 years, he is likely to have died from other causes - due to the high competing mortality rates in older men. We submit that it is unambigious that the trade-off between benefits and harms from screening younger men is quite different than for older men.

The UK, like many other high-income countries, has an opportunistic screening policy. This is suboptimal because it has, effectively, led to screening in men older than 70 years.[23, 24] PSA testing rates in the UK in 2018 were three times higher for men aged 70-79 than 50-59 years.[15, 25] There is also a policy of PSA testing in older men with erectile dysfunction, hematuria, and lower urinary tract symptoms (LUTS). The problem with this is that these are not symptoms of localised prostate cancer or associated with increased risk of high-grade prostate cancer,[26] and they are extremely common in older men.[27] Thus ‘symptomatic’ testing is effectively asymptomatic screening, also likely to result in overdiagnosis. The result of this and the opportunistic screening policy is that the number of older men receiving PSA tests has risen substantially in recent years.[6, 28] While there is some guidance on PSA testing for men aged 80 years or older,[29] ensuring informed decision making is a challenge for primary care general practitioners. Men have access to conflicting information on harms and benefits such as from different websites, which is also reflected in differing interpretations of the evidence from various guideline developers.[3, 4] Some tools are available to support decision making focused on checking risk from prostate cancer; but none appear to evaluate risk of overdiagnosis based on a man’s age. These could help ensure men in shared decision making contexts are adequately informed about risk of harm due to overdiagnosis from PSA testing in relation to their age and other life expectancy risk factors (see data availability section for an example app prototype).

Our analysis provides indirect support to the hypothesis that opportunistic testing, as currently implemented in the UK, is worse than a policy of either organised risk-based screening,[3] or no access to opportunistic PSA testing at all.[30] However, it is insufficient to determine what type of organised screening, if any, should be recommended in its place. A more comprehensive evaluation is needed for this, including to estimate the impact of substantially earlier diagnosis from screening younger men on their quality of life, and overall cost-effectiveness. Other work has considered these issues, including to inform the recent UK NSC draft statement on prostate cancer screening.[7] The findings from our analysis provide a means to evaluate the face validity of overdiagnosis projections from this model. The model projected overdiagnosis from a one-off screen to be 28% for a 50 year old, increasing to 82% for a 70 year old;[7] and an associated infographic gave a headline figure of 71% (20/28) for screening men aged 50-60 years.[31] Based on our analysis these are substantial overestimates. We also note that the model evaluated a policy of organised screening on top of opportunistic testing, which is a policy few would advocate irrespective of modeling. For example, the analysis did not consider that an organised program could reduce overdiagnosis compared with opportunistic testing, by ending the availability of PSA testing through primary care for asymptomatic older men. Therefore, in our view, the model needs further work before it may be considered sufficiently reliable to project the impact of organised-screening regimens on overdiagnosis relative to the current opportunistic testing policy. Further data are also needed to evaluate the impact of harms from overdiagnosis in the modern era. This includes on the use of active surveillance,[32] and focal therapy[33]. Ongoing trials are anticipated to provide such evidence to help further guide policymakers.

## Data Availability

All data used and analysis code are available online at https://github.com/brentnall/pca-overdx

## Acknowledgment

We are grateful to Stephen Duffy for comments on an earlier version of this report.

## Data Availability Statement

All data used and analysis code are available at https://github.com/brentnall/pca-overdx, and an overdiagnosis calculator app prototype.

